# Using Media to Promote Public Awareness of Early Detection of Kaposi’s Sarcoma in Africa

**DOI:** 10.1101/2019.12.11.19013649

**Authors:** Miriam Laker-Oketta, Lisa Butler, Philippa Kadama-Makanga, Robert Inglis, Megan Wenger, Edward Katongole-Mbidde, Toby Maurer, Andrew Kambugu, Jeffrey Martin

## Abstract

**Background:** Despite its hallmark cutaneous presentation, most Kaposi’s sarcoma (KS) in Africa is diagnosed too late for effective treatment. Early diagnosis will only be achievable if patients with KS present earlier for care. We hypothesized that public awareness about KS can be enhanced through exposure to common media.

**Methods:** We developed educational messages regarding early detection of KS for the general African public portraying a three-part theme: “Look” (regularly examine one’s skin/mouth); “Show” (bring to the attention of a healthcare provider any skin/mouth changes); and “Test” (ask for a biopsy for definitive diagnosis). We packaged the messages in three common media forms (comic strips, radio, and video) and tested their effect on increasing KS awareness among adults attending markets in Uganda. Participants were randomized to a single exposure to one of the media and evaluated for change in KS-related knowledge and attitudes.

**Results:** Among 420 participants, media exposure resulted in increased ability to identify KS (from 0.95% pre-test to 46% post-test); awareness that anyone is at risk for KS (29% to 50%); belief that they may be at risk (63% to 76%); and knowledge that definitive diagnosis requires biopsy (23% to 51%) (all p<0.001). Most participants (96%) found the media culturally appropriate.

**Conclusion:** Exposure to media featuring a theme of “Look”, “Show”, “Test” resulted in changes in knowledge and attitudes concerning KS among the general public in Uganda. High incidence and poor survival of KS in Africa are an impetus to further evaluate these media, which are freely available online.

## Introduction

A result of the intersection between the HIV epidemic and the endemic presence of Kaposi sarcoma-associated herpesvirus (KSHV) infection, sub-Saharan Africa accounts for over 80% of incident Kaposi’s sarcoma (KS) worldwide (1). In East Africa, for example, KS is the most frequently reported cancer among men and third in women. In spite of the incidence of KS in sub-Saharan Africa and its hallmark presentation on the skin and visible mucous membranes (i.e., in areas where it theoretically can be detected in an early stage), more than 80% of all KS in sub-Saharan Africa is classified as late-stage at the time of diagnosis (2-4). Like many cancers, KS diagnosed in advanced stages has worse survival than early diagnosis (4, 5). Probably more than most cancers, however, detecting KS early in sub-Saharan Africa could likely largely control the condition. This is because early KS can often be treated with widely available drugs — the antiretroviral therapy (ART) that is used for HIV infection (6). Late-stage KS, in contrast, is best treated with liposomal anthracyclines or paclitaxel (6), which, because of cost, are scarcely available.

Because HIV-infected individuals are at particular risk for KS, a natural target to consider for interventions to enhance early detection of KS in Africa are the numerous primary care facilities that have arisen to enable the roll-out of ART. Recent work from Zimbabwe, however, which provided training regarding early KS detection to frontline clinicians at HIV-dedicated primary care clinics, demonstrated that training alone may be insufficient. In this research, providing training increased clinician knowledge but had no effect on stage of patients’ KS at diagnosis (7). In retrospect, this finding is unsurprising. If healthcare providers are primarily only faced with KS in its late stages, they will not have the opportunity to make earlier diagnoses. Furthermore, if patients who develop KS are unaware of what it is and the importance of seeking immediate care, it will continue to be difficult to get patients to present to healthcare providers in the early stages of KS. Although general public awareness has not been studied, we earlier found that only 7% of HIV-infected patients — the group with the most at stake regarding KS — had heard of the condition (8).

To address the gap in public knowledge about KS in sub-Saharan Africa and ultimately facilitate early detection, we endeavored to create an educational message regarding KS and the importance of early diagnosis that could be disseminated in the community via common media formats. We subsequently tested whether exposure to the media forms was able to change knowledge and attitudes regarding KS among adults in the general community.

## Materials and Methods

### Overall design

Using community-engaged research, we developed an educational message regarding KS and its early detection and incorporated this message into three common media forms. To determine whether the message and media forms influenced knowledge and attitudes concerning KS among members of the general public, we then randomly assigned exposure to the three media forms to adults attending rural and urban commercial markets in Uganda. Lastly, we assessed knowledge and attitudes concerning KS with pre- and post-media tests.

### Development of media

Our objective was to develop a scientifically sound and culturally appropriate educational message about KS and its early detection. We, also, aimed to determine if common media forms could be used to disseminate the message. We worked with an international health science communication consultant (Jive Media Africa, South Africa) and used community-engaged research. Community engagement involved soliciting input from key stakeholders, at several different stages, including a) patients with KS who had survived the cancer (diagnosed in early stages and still alive 7 to 10 years later) and persons living with HIV infection; b) healthcare practitioners including primary healthcare providers, community health workers, and traditional health practitioners; c) community leaders and members of the general public; and d) local media groups experienced with health message dissemination in Uganda.

We developed an educational message with a three-part theme: “Look”, indicating to perform skin and mouth examination, either by one’s self or via a partner, to identify any new or unusual skin/mouth lesions; “Show”, meaning to bring any suspicious skin/mouth lesions to the attention of a healthcare provider; and “Test”, denoting to ask healthcare providers to perform biopsies for suspicious lesions. We incorporated this “Look”; “Show”; “Test” theme into storylines featuring fictional and real-life (healthcare providers and patients with KS) characters finding abnormal skin lesions, presenting the lesions to healthcare providers, receiving a skin punch biopsy for diagnosis, and initiating treatment for KS. The message was crafted drawing upon the concepts of the Information–Motivation–Behavioral skills model (IMB model) (9). To disseminate the message, we chose three common media forms: paper-based multi-panel comic strips (Figure 1); radio (i.e., audio) vignettes (https://soundcloud.com/kaposis_sarcoma); and a short (10-minute) video documentary (https://vimeo.com/224920054). All media were created in English and the four most commonly spoken local languages (Luganda, Luo, Runyakitara, and Kiswahili).

**Figure 1.**
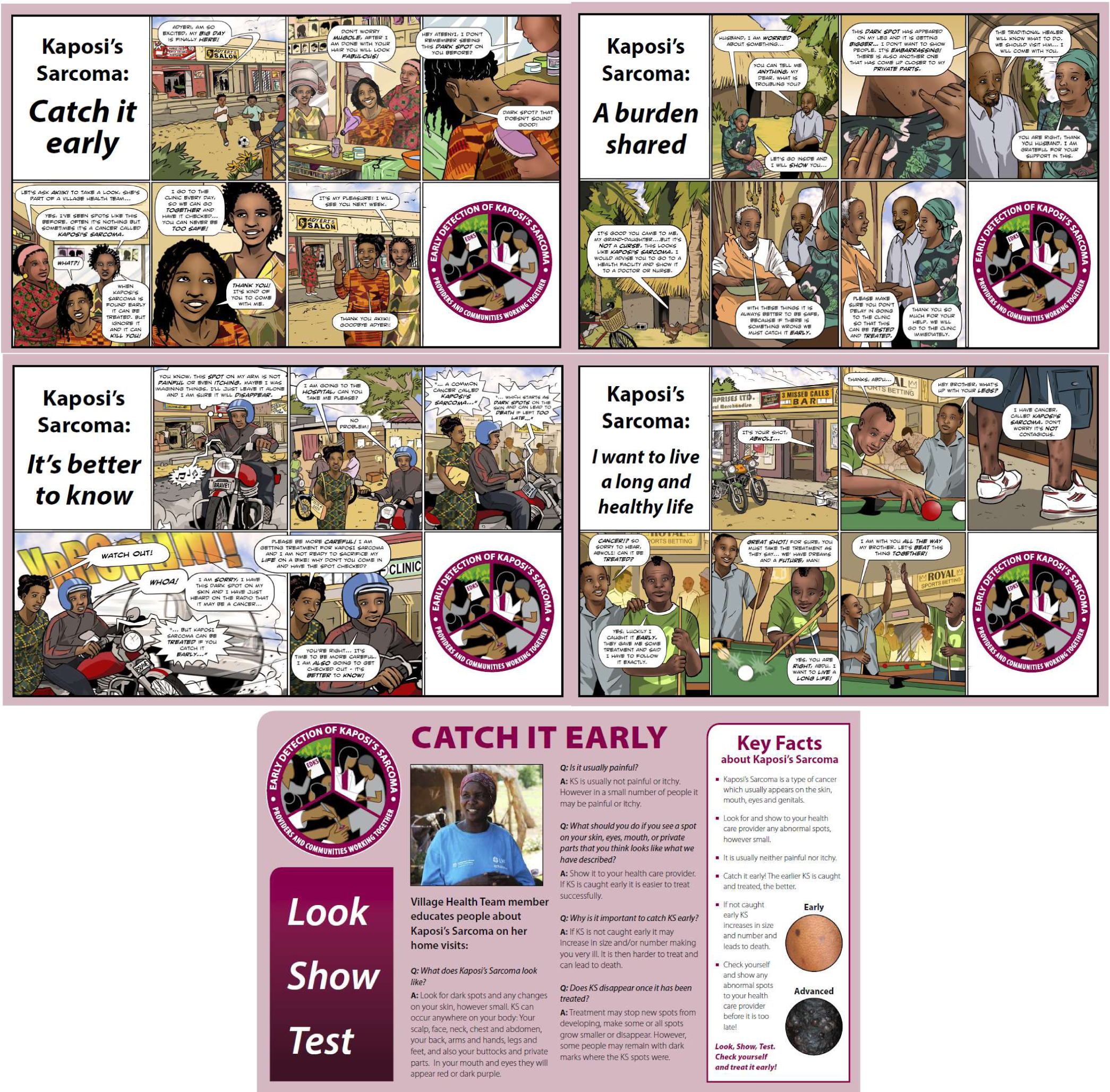
Comic strips, one of three common media forms regarding KS awareness and early detection developed and evaluated among a community-based sample of Ugandan adults.

### Evaluation of media

#### Study population

The target population for our evaluation of the three media forms was the general Ugandan adult population aged at least 18 years. As an accessible population, we choose to sample participants from commercial markets. Markets in Uganda are concentrations of multiple open-air structures where food (especially perishables) and everyday household items are traded. Each market serves a given approximate geographic area. We chose markets from one predominantly rural district (Hoima, in western Uganda, defined as 82% rural (10)) and the two most populated urban districts, Kampala (the capital city) and Wakiso (which surrounds Kampala). In each area, first, we randomly selected administrative units. We then used local government records to locate the markets within these administrative units and randomly chose markets to include in the media evaluation. At each selected market, no advertising or announcement was made prior to the day of media testing. On the media evaluation day, we used the markets’ public-address systems as well as one-on-one random outreach, by local community mobilizers, to inform market attendees about our presence and invite them to visit our booth to learn more. The initial explanation described our work as a study about health. Those who responded to the invitation to visit our booth were given more information about what they were being asked to do. We did not use the terms HIV/AIDS, Kaposi’s sarcoma, cancer, or ART. We enrolled consecutive interested volunteers, stratified by gender, seeking an equal number of men and women. Enrollment continued until we had attained the requisite participant numbers for both sexes. The project received institutional review board approval (National HIV/AIDS Research Committee #138), and participants provided verbal informed consent.

#### Exposure to media

We randomly assigned participants, stratified by gender, to one of the three media types. The first was the comics which we printed on newspaper-sized hard stock cards. The second was the video documentary which we showed on hand-held electronic tablets. The third, the radio vignettes, were administered through hand-held radios. Because we wanted to simulate a single exposure to the media, participants were only allowed to possess the media for a limited amount of time (ten minutes each for the comics and radio vignettes and 15 minutes for the video), following which they were immediately administered a post-test. Illiterate participants assigned to comics had a research team member read each story once while the participant viewed the corresponding pictures. Any questions about the material were deferred until after testing was completed.

#### Main outcome assessment

Our main objective was to describe the change in knowledge and attitudes concerning KS as a result of being exposed to the media. We determined within-person change by comparing responses on an interviewer-administered test completed prior to the exposure to the media to the responses on the same test administered immediately after the exposure. The test questions were informed by the concepts of the IMB model. They covered visual recognition of KS (by showing participants pictures of KS lesions); knowledge of the causative agent and casual behavioral factors; self-risk assessment; and health-seeking attitudes regarding KS.

#### Other variables

The interviewer-administered pre-test also obtained information on age, sex, education and literacy level, residence, income, and self-reported HIV status.

#### Statistical analysis

Each item on the questionnaire had a single best specific response according to the messages covered by the media. For some of the items, additional correct (typically less specific) responses were also possible. For each item, we determined within-participant change. We determined the percentage of participants whose responses changed from incorrect on the pre-test to correct on the post-test; percentage of participants whose responses changed from correct to incorrect, and percentage whose responses did not change. We performed this analysis for the single best response and, separately, for all possible correct responses. Statistical significance of the within-participant change was determined with McNemar’s test. We also evaluated the influence of several factors (age, sex, education level, residence, and self-reported HIV status) on change in KS knowledge and attitudes. The outcome was a change from an incorrect response on the pre-test to a correct response on the post-test; this was coded as “1”. All those whose responses did not change or changed in the negative direction were coded as “0”. We used a directed acyclic graph (11, 12) to depict background knowledge and inform variable selection in the multivariable logistic regression model (Figure 2). All analyses were performed with Stata version 13.1 (Stata Corp., College Station, Texas).

**Figure 2.**
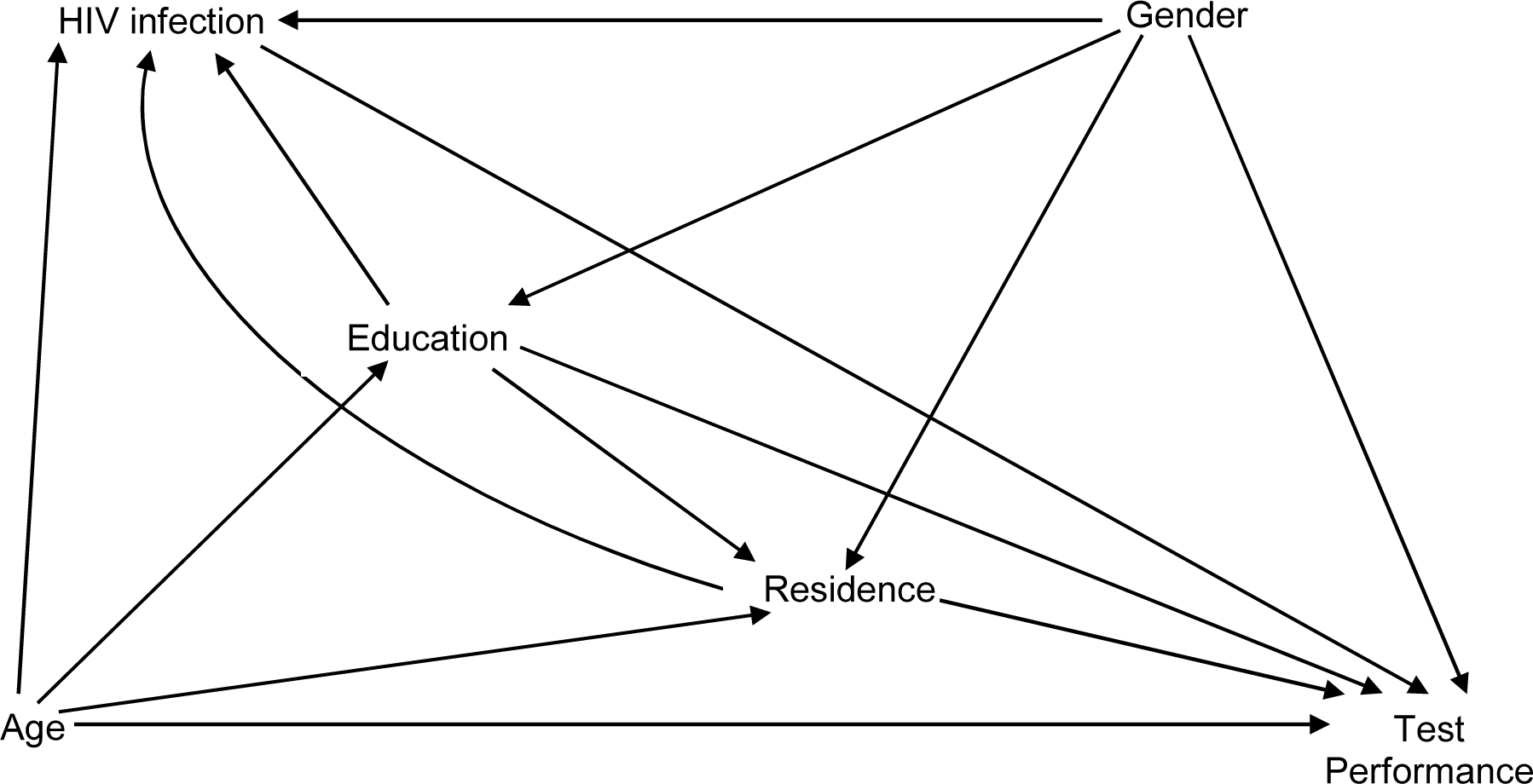
Directed acyclic graph showing possible relationships between potential causal determinants and performance on a written evaluation following exposure to common media forms regarding KS awareness and early detection among community-based adults in Uganda.

## Results

### Characteristics of the study population

From February 2017 to January 2018, we examined 420 adults from urban (n=237) and rural (n=183) Uganda. The median age was 30 years (interquartile range: 24 to 39), and, by design, 50% were women (Table 1). Half of the participants had attained education beyond primary school level. Literacy, measured by the ability of participants to comfortably read grade 5-level writing in a language of their choice, was 67% (Overall literacy in Uganda is 72% (10)). HIV prevalence, based on self-report, was 5.5% which is equivalent to the national prevalence of 6.2% (13).

**Table 1.** Characteristics of a community-based of Ugandan adults enrolled in a study of exposure to common media forms regarding KS awareness and early detection, overall and by media form.

| Characteristic | Comics<br>(n=140) | Video<br>(n=140) | Radio Vignettes<br>(n=140) | All<br>(N=420) |
| --- | --- | --- | --- | --- |
| **Age, in years\*** | 29 (23 to 38) | 30 (25 to 40) | 31 (25 to 40) | 30 (24 to 39) |
| **Female gender** | 50% | 51% | 50% | 50% |
| **Residence** |  |  |  |  |
| Urban | 56% | 57% | 56% | 56% |
| Rural | 44% | 43% | 44% | 44% |
| **Marital status** |  |  |  |  |
| Married | 56% | 51% | 59% | 55% |
| Widowed | 4.0% | 4.0% | 6.0% | 5.0% |
| Divorced | 17% | 18% | 16% | 17% |
| Never married | 23% | 27% | 19% | 23% |
| **Education level** |  |  |  |  |
| None/Primary | 51% | 50% | 49% | 50% |
| Lower Secondary | 33% | 33% | 31% | 33% |
| Higher Secondary/Tertiary | 16% | 17% | 20% | 17% |
| **Literacy†** | 64% | 66% | 71% | 67% |
| **Annual income, in US dollars\*\*** | 833<br>(500 to 1667) | 833<br>(417 to 1500) | 1000<br>(483 to 1583) | 833<br>(500 to 1667) |
| **HIV-infected, via self-report** | 3.6% | 6.5% | 6.5% | 5.5% |

\* median (interquartile range).
† Participants were considered literate if they were able to read three sentences in Grade 5-level writing in the language of their choice.
‡ Exchange rate: 1 US Dollar = 3600 Uganda Shillings

### Baseline awareness of KS and early detection

Before participants were exposed to the media materials, we assessed their knowledge and attitudes regarding KS. When shown colored photographs of cutaneous and oral macular, plaque, and nodular KS in different anatomical locations, only 0.95% of participants correctly identified the lesions as KS (Table 2). While 29% of participants correctly believed that anyone could develop KS, 63% considered themselves to be at risk for it. The most common four factors participants thought made them at risk to develop KS were: all people were at risk (22%); a belief that KS is contagious and that they are at risk by sharing fomites or unknowingly coming into physical contact with people with KS (16%); poverty (11%); and being sexually active (9.0%) (Figure 3). Even though 60% said they would go immediately to a healthcare provider in the event of development of lesions like those shown in the photographs on their skin or mouth, only 23% of participants appreciated that a biopsy (i.e., some technical examination of the lesion) is necessary to correctly identify the condition.

**Table 2.** Change in KS knowledge and attitudes following exposure to common media forms, overall and by media type, among a community-based sample of adults in Uganda.

| Question | Comics<br>(n=140) |  |  | Video<br>(n=140) |  |  | Radio Vignettes<br>(n=140) |  |  | All participants<br>(N=420) |  |  |
| --- | --- | --- | --- | --- | --- | --- | --- | --- | --- | --- | --- | --- |
|  | Pre-test | Post-test | Change | Pre-test | Post-test | Change | Pre-test | Post-test | Change | Pre-test<br>(95% CI) | Post-test<br>(95% CI) | Change |
| **Shown a picture of cutaneous KS: What do you think this condition is?** |  |  |  |  |  |  |  |  |  |  |  |  |
| “Kaposi’s sarcoma” | 0.71% †<br>(0.02% - 3.9%) | 38%<br>(30% - 46%) | 37%‡<br>0.0%§<br>0.7%¶<br>62%**<br>p<0.001†† | 0.71%<br>(0.02% - 3.9%) | 52%<br>(44% - 61%) | 51%<br>0.0%<br>0.7%<br>49%<br>p<0.001 | 1.4%<br>(0.17% - 5.1%) | 47%<br>(39% - 56%) | 46%<br>0.0%<br>1.4%<br>53%<br>p<0.001 | 0.95%<br>(0.3%- 2.4%) | 46%<br>(41%-51%) | 45%<br>0.0%<br>1.0%<br>55%<br>p<0.001 |
| “Kaposi’s sarcoma”, or “skin cancer”, or “unspecified cancer” | 44%<br>(35% - 52%) | 86%<br>(80% - 92%) | 46%<br>2.9%<br>41%<br>11%<br>p<0.001 | 40%<br>(32% - 49%) | 85%<br>(78% - 90%) | 46%<br>1.4%<br>39%<br>14%<br>p<0.001 | 45%<br>(37% - 54%) | 84%<br>(77% - 90%) | 42%<br>2.9%<br>42%<br>13%<br>p<0.001 | 43%<br>(38%-48%) | 85%<br>(81%-88%) | 45%<br>2.4%<br>41%<br>12%<br>p<0.001 |
| **What do you think causes it?** |  |  |  |  |  |  |  |  |  |  |  |  |
| “HHV-8/KSHV” or “a microorganism” | 3.6%<br>(1.2% - 8.1%) | 3.6%<br>(1.2% - 8.1%) | 2.1%<br>2.1%<br>1.4%<br>94%<br>p=1.00 | 5.0%<br>(2.0% - 10%) | 8.6%<br>(4.5% - 14%) | 6.4%<br>2.9%<br>2.1%<br>89%<br>p=0.27 | 2.1%<br>(0.4% - 6.1%) | 5.7%<br>(2.5% - 11%) | 5.0%<br>1.4%<br>0.7%<br>93%<br>p=0.18 | 3.6%<br>(1.8%-5.4%) | 6.0%<br>(3.7%-8.2%) | 4.5%<br>2.1%<br>1.4%<br>92%<br>p=0.09 |
| “HHV-8/KSHV”, “HIV”, or “a microorganism” | 9.3%<br>(5% - 15%) | 11%<br>(6.1% - 17%) | 4.3%<br>2.9%<br>6.4%<br>86%<br>p=0.75 | 8.6%<br>(4.5% - 14%) | 11%<br>(6.1% - 17%) | 7.9%<br>5.7%<br>2.9%<br>84%<br>p=0.65 | 11%<br>(6.1% - 17%) | 16%<br>(11% - 24%) | 11%<br>5.0%<br>5.7%<br>79%<br>p=0.13 | 9.5%<br>(6.7%-12%) | 13%<br>(9.4%-16%) | 7.6%<br>4.5%<br>5.0%<br>82.9%<br>p=0.07 |
| **Who is at risk for this condition?** |  |  |  |  |  |  |  |  |  |  |  |  |
| “All people” | 29%<br>(22% - 38%) | 44%<br>(35% - 52%) | 26%<br>12%<br>17%<br>44%<br>p=0.01 | 28%<br>(21% - 37%) | 58%<br>(49% - 66%) | 37%<br>7.1%<br>21%<br>35%<br>p<0.001 | 29%<br>(21% - 37%) | 47%<br>(39% - 56%) | 29%<br>10%<br>19%<br>43%<br>p=0.001 | 29%<br>(24%-33%) | 50%<br>(45%-54%) | 31%<br>9.8%<br>19%<br>40%<br>p<0.001 |
| “HIV infected” | 5.7%<br>(2.5% - 11%) | 5.0%<br>(2.0% - 10%) | 5.0%<br>5.7%<br>0.0%<br>89%<br>p=1.00 | 1.4%<br>(0.17% - 5.1%) | 6.4%<br>(3.0% - 12%) | 5.7%<br>0.7%<br>0.7%<br>93%<br>p=0.04 | 5.0%<br>(2.0% - 10%) | 15%<br>(9.5% - 22%) | 14%<br>3.6%<br>1.4%<br>81%<br>p=0.01 | 4.0%<br>(2.4% - 6.4%) | 8.8%<br>(6.3% - 12%) | 8.1%<br>3.3%<br>0.7%<br>88%<br>p=0.001 |
| "All people" or<br>"HIV-infected" | 35%<br>(27% - 44%) | 47%<br>(39% - 56%) | 26%<br>14%<br>21%<br>39%<br>p=0.03 | 29%<br>(22% - 38%) | 61%<br>(53% - 70%) | 39%<br>6.4%<br>23%<br>32%<br>p<0.001 | 34%<br>(26% - 42%) | 59%<br>(50% - 67%) | 32%<br>7.1%<br>26%<br>34%<br>p<0.001 | 33%<br>(28%-37%) | 56%<br>(51%-60%) | 32%<br>9.0%<br>24%<br>35%<br>p<0.001 |
| **Would you consider yourself to be at risk?** |  |  |  |  |  |  |  |  |  |  |  |  |
| "Yes" | 62%<br>(54% - 70%) | 75%<br>(67% - 82%) | 21%<br>7.9%<br>54%<br>17%<br>p=0.004 | 62%<br>(53% - 70%) | 79%<br>(71% - 85%) | 22%<br>4.3%<br>56%<br>16%<br>p<0.001 | 66%<br>(57% - 74%) | 75%<br>(67% - 82%) | 15%<br>5.7%<br>59%<br>19%<br>p=0.02 | 63%<br>(58%-68%) | 76%<br>(72%-80%) | 19%<br>6.0%<br>57%<br>18%<br>p<0.001 |
| **What would you do if you began to see similar lesions on your skin?** |  |  |  |  |  |  |  |  |  |  |  |  |
| "Go to biomedical<br>provider" or<br>"get a biopsy" | 59%<br>(50% - 67%) | 96%<br>(92% - 99%) | 38%<br>0.0%<br>59%<br>3.6%<br>p<0.001 | 57%<br>(49% - 65%) | 96%<br>(91% - 98%) | 39%<br>0.0%<br>57%<br>4.3%<br>p<0.001 | 63%<br>(54% - 71%) | 96%<br>(91% - 98%) | 35%<br>2.1%<br>61%<br>2.1%<br>p<0.001 | 60%<br>(55%-64%) | 96%<br>(94%-98%) | 37%<br>0.7%<br>59%<br>3.3%<br>p<0.001 |
| **How do healthcare providers figure out or find out what this condition is to give you the right treatment? ††** |  |  |  |  |  |  |  |  |  |  |  |  |
| "Perform a biopsy" | 20%<br>(9.1% - 36%) | 35%<br>(21% - 52%) | 20%<br>5.0%<br>15%<br>60%<br>p=0.11 | 25%<br>(13% - 41%) | 73%<br>(56% - 85%) | 53%<br>5.0%<br>20%<br>23%<br>p<0.001 | 23%<br>(11% - 39%) | 46%<br>(30% - 63%) | 28%<br>5.1%<br>18%<br>48%<br>p=0.02 | 23%<br>(15%-31%) | 51%<br>(42%-61%) | 34%<br>5.0%<br>18%<br>44%<br>p<0.001 |
| "Perform a biopsy"<br>or "skin exam" | 45%<br>(29% - 62%) | 63%<br>(46% - 77%) | 28%<br>10%<br>35%<br>28%<br>p=0.12 | 43%<br>(27% - 59%) | 85%<br>(70% - 94%) | 45%<br>2.5%<br>40%<br>13%<br>p<0.001 | 44%<br>(28% - 60%) | 56%<br>(40% - 72%) | 27%<br>13%<br>31%<br>31%<br>p=0.30 | 44%<br>(35%-53%) | 68%<br>(59%-76%) | 33%<br>8.4%<br>35%<br>24%<br>p<0.001 |

\*Correct response refers to the current best scientific evidence regarding the question. This is the information that was promulgated in each of the three common media forms. It is the first response listed below the question. For some questions, we also considered responses closely related to the correct response; these are listed in the second and third rows. In rows which include 'or', we considered either of the listed responses.
† % (95% confidence intervals) of participants who provided the response.
‡ % of participants who had "incorrect" response on pre-test and the "correct" index response on post-test.
§ % of participants who had index "correct" response on pre-test and "incorrect" response on post-test.
¶ % of participants who had "correct" responses on both pre-test and post-test.
\*\* % of participants who had "incorrect" responses on both pre-test and post-test.
†† p value for within-subject change from McNemar's test.
†† Limited to N=119 participants: 40 randomized to comics, 40 to video, and 39 to radio vignettes. The question was incorporated in a later version of the pre- and post-test.

**Figure 3.**
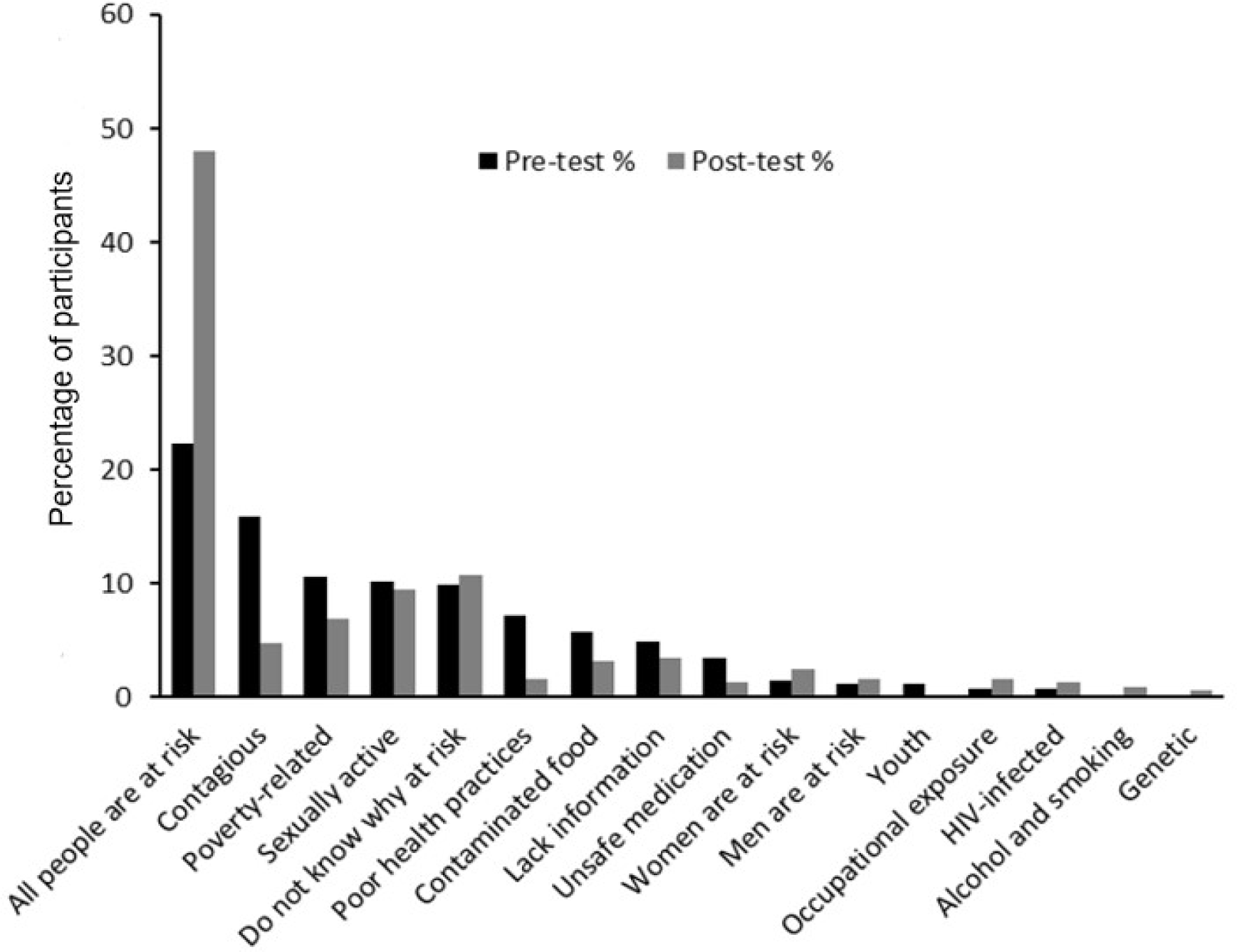
Explanations cited for being at risk to develop KS by a community-based sample of Ugandan adults who thought themselves to be at risk for KS, prior to (pre-test N= 265) and following (post-test N= 319) exposure to one of three media forms regarding KS awareness and early detection.

### Changes in knowledge and attitude after exposure to media

Participants were re-tested with the same set of questions immediately after being exposed, via random assignment, to one of the three media forms. There was a significant increase in the percentage of participants responding correctly to all but one of the questions posed (Table 2). The largest overall correct change in performance was in the ability of participants to specifically name KS as the lesions depicted in the photographs; 46% named KS after media exposure (an absolute increase of +45% compared to pre-media knowledge). This was also the only question where, across all media types, none of the respondents who had the correct response at pre-test gave an incorrect response at post-test. Approximately one-third of all participants correctly changed their responses to the questions asking about who is at risk for KS (+31% changed to “all people”); what they would do if they saw the lesions themselves on their skin (+37% changed to “go to a biomedical provider or get a biopsy”); and what is it that a healthcare provider would do to make the diagnosis of KS (+34% changed to “perform a biopsy”). In general, there were significant increases in correct responses in each of the comics, video documentary, and radio vignette-exposed groups. Of note, we also observed participants changing their response from a correct to an incorrect answer although this was typically of considerably lower magnitude than the other direction.

The smallest but yet still significant change in test performance was in the question which asked the participants if they considered themselves at risk for KS. After media exposure, 19% more participants considered themselves at risk for KS. The rationale the participants gave for believing themselves to be at risk also changed in comparison to the pre-media evaluation (Figure 3). Specifically, in the post-test, citing that all people were at risk was again the most common reason given for the participant believing him/herself at risk, but this was now claimed by 50% of those believing they were at risk. A belief that KS is contagious decreased to only being present post-test in 5.0% of participants claiming to be at risk.

Exposure to the media did not, overall, result in any significant change in participants correctly identifying that KS is caused by a microorganism. This remained the case even when HIV was considered an acceptable response as a cause of KS along with the most specific correct answer (human herpesvirus 8/Kaposi’s sarcoma-associated herpesvirus (HHV-8/KSHV) and “microorganism”.

### Determinants of change in knowledge and attitude after exposure to media

We explored the relationship between various participant characteristics and ability, after media exposure, to provide correct responses on the post-test. We evaluated these relationships in the context of three test questions: ability to identify and cite KS by name in response to photographs; knowledge of who is at risk for KS; and recognition that any suspicious skin or mouth changes should be presented immediately to a biomedical healthcare provider (Table 3). In adjusted analyses, only two of the variables, current residence, and education level attained were associated with improved test performance. While participants residing in urban areas had 49% (95% CI: 22% to 67%; p = 0.002) lower odds of correctly identifying KS from photographs compared to those residing in rural areas, those in urban areas had higher odds of correct responses in the other two questions. Having at least secondary education had higher odds of correctly naming KS compared to having only primary or no formal education. Notably, education level showed no strong evidence of an association with performance on the other two questions.

**Table 3.** Evaluation of potential causal determinants of performance on a written evaluation following exposure to common media forms regarding KS awareness and early detection among a community-based sample of Ugandan adults.

| Characteristic | Question<br>“Correct Response” |  |  |
| --- | --- | --- | --- |
|  | Shown a picture of cutaneous KS:<br>What do you think this condition is? | Who is at risk for this<br>condition? | What would you do if you began to see<br>similar lesions on your skin? |
|  | “Kaposi’s sarcoma” | “All people or HIV-infected” | “Get a biopsy or go to a biomedical provider” |
|  | OR (95% CI; p value) | OR (95% CI; p value) | OR (95% CI; p value) |
| **Age**, per 10-year increase* | 0.86 (0.71 to 1.0; p = 0.13) | 1.1 (0.91 to 1.3; p = 0.31) | 0.96 (0.72 to 1.2; p = 0.72) |
| **Gender**<sup>†</sup> |  |  |  |
| Men | Ref. | Ref. | Ref. |
| Women | 1.4 (0.94 to 2.3; p = 0.95) | 0.89 (0.58 to 1.4; p = 0.60) | 1.2 (0.72 to 2.1; p = 0.44) |
| **Residence**<sup>‡</sup> |  |  |  |
| Rural | Ref. | Ref. | Ref. |
| Urban | 0.51 (0.33 to 0.78; p = 0.002) | 1.9 (1.2 to 2.9; p = 0.005) | 63 (25 to 163; p < 0.001) |
| **Education level**<sup>§</sup> |  |  |  |
| None or primary | Ref. | Ref. | Ref. |
| Lower secondary | 1.9 (1.2 to 3.1; p = 0.006) | 1.2 (0.75 to 1.9; p = 0.44) | 0.98 (0.54 to 1.8; p = 0.95) |
| Higher secondary or tertiary | 5.1 (2.8 to 9.4; p < 0.001) | 0.87 (0.42 to 1.6) p = 0.66) | 1.2 (0.61 to 2.5; p = 0.55) |
| **HIV infection status**<sup>¶</sup> |  |  |  |
| Uninfected | Ref. | Ref. | Ref. |
| HIV-infected | 0.54 (0.21 to 1.4; p = 0.21) | 1.8 (0.75 to 4.3; p = 0.19) | 0.39 (0.14 to 1.1; p = 0.07) |
| Never tested | 0.51 (0.19 to 1.3; p = 0.17) | 1.3 (0.50 to 3.1; p = 0.63) | 0.29 (0.08 to 1.0; p = 0.05) |

\* adjusted for gender, residence, education level, and HIV status
† adjusted for age, residence, education level, and HIV status
‡ adjusted for age, gender, education level, and HIV status
§ adjusted for age, gender, residence, and HIV status
¶ adjusted for age, gender, residence, and education

### Participants’ assessment of the media materials

When asked about their opinions about the suitability of the educational materials, 93% of participants were satisfied with the media content and offered no suggestions for improvement. Of the 31 (7.4%) participants who suggested some adjustments, 19% believed that including more photographs of KS lesions would help reinforce the messages, and 29% requested more information on the causative agent of KS (Table 4). Only 16 (3.8%) participants reported finding aspects inappropriate for public consumption.

**Table 4.** Open-ended responses regarding suggestions for improvement and perceived inappropriate content amongst a community-based sample of Ugandan adults exposed to common media forms regarding KS awareness and early detection

| Question and Response | No. (%) of Participants<br>Endorsing<br>(N=420) |
| --- | --- |
| **What do you think should be added to the media to help you or others understand it better?** |  |
| More information on the cause of KS | 9 (2.1%) |
| More KS images | 6 (1.4%) |
| More information encouraging health-seeking with KS | 4 (0.95%) |
| KS prevention | 4 (0.95%) |
| Testimonies and before and after photographs of survivors | 2 (0.48%) |
| KS transmission | 2 (0.48%) |
| KS progression | 1 (0.24%) |
| HIV-specific information and images | 1 (0.24%) |
| Information on other cancers | 1 (0.24%) |
| KS in children | 1 (0.24%) |
| **Is there anything that you or people in your community might find offensive/inappropriate/disturbing in the way the characters look, their actions, or the words they are using?** |  |
| Something offensive/inappropriate/disturbing | 16 (3.8%) |
| **What might be offensive/inappropriate/disturbing, and why?** |  |
| People seeking help from traditional healers was troubling | 4 (0.95%) |
| Images of KS lesions were distressing | 4 (0.95%) |
| Scene where woman exposes her inner thigh to show lesions was embarrassing | 3 (0.71%) |
| Could not recall the exact scene | 3 (0.71%) |
| Scene depicting the skin punch biopsy process was disturbing | 1 (0.24%) |
| When a motorcycle accident almost occurred | 1 (0.24%) |

## Discussion

Despite the possibility of remission when diagnosed at an early stage, KS in sub-Saharan Africa is too often detected at advanced disease states for which available treatment is ineffective. The irony is exacerbated by most KS presenting on the skin or mouth, areas in which abnormalities should be readily identified and brought to medical attention. In a community-based sample of Ugandan adults, we found — at least in part explaining why advanced stage presentation is so frequent — that knowledge about KS and its early detection was limited. To begin to remedy this, we developed an educational message about KS targeted to the public. The themes intended to educate the public about the importance of examining their skin (“Look”); timely presentation to a healthcare provider if abnormalities are found (“Show”), and asking providers to perform biopsies for definitive diagnosis (“Test”). To disseminate the message, we incorporated it into three common media forms: comics, radio vignettes, and a video documentary. We demonstrated that exposure to these media increased knowledge and changed attitudes about KS and its early detection. The messages and modes of delivery were acceptable to the public, thus providing a promising foundation for future comprehensive educational efforts. To our knowledge, this is the first formal effort to begin to educate the public in Africa about KS.

We found very low knowledge regarding KS in our participants prior to exposing them to the media materials. Almost no participant could recognize and cite “Kaposi sarcoma”, and less than 50% thought of it as cancer. There was also no local terminology or name used to refer to KS. This low level of specific knowledge in the population has been documented for other cancers in sub-Saharan Africa (14-16). Although more than half of the participants believed themselves to be at risk for KS, the majority of risk factors mentioned were incorrect. These explanations for risk included potential for contracting KS from another person with KS, poverty, and sexual contact, all of which are commonly held beliefs about how other cancers develop in Africa (17). Although seemingly independent of the ability to name KS or its causal determinants, we also found that a large fraction of participants were unaware of the need to seek immediate attention from a biomedical provider if they developed lesions resembling KS. We did not probe why participants deemed it unimportant to seek immediate medical attention but suspect it is similar to what has been reported for other cancers in the region. That is, for people who are concerned about the meaning of such lesions, failure to seek care from a biomedical provider could be because of access problems including but not limited to transportation, competing work and social responsibilities, reliance on alternative healthcare (e.g., traditional healers), and use of non-prescription medicines and natural remedies (18, 19). For others, if lesions are not accompanied by any other symptoms (e.g., pain) or do not rapidly change, there simply may be insufficient knowledge that new spots on one’s skin may portend a serious condition. Indeed, studies of late presentation of other cancers in Africa have shown that the choice to seek help often only arises when symptoms persist or worsen (18-22). Finally, not only was there substantial disregard for immediate medical attention if lesions resembling KS developed, there was also scant awareness of the need for biopsy to establish a definitive diagnosis.

After a single setting exposure to one of the three media forms, participants exhibited significant increases in their ability to identify and name KS, recognize that everyone is at risk for KS, state that they would seek attention from a biomedical provider if they developed lesions resembling KS, and know that a biopsy is needed to establish a diagnosis. We are unaware of similar educational efforts targeted at the general public to enhance knowledge about KS. Community-targeted educational efforts have, however, been implemented for breast and cervical cancer in Africa with demonstrable increases in knowledge (23-25). While increase in KS knowledge is not a guarantee of change in behavior, and despite absence of evidence of analogous clinical impact in other cancers in sub-Saharan Africa, public education is known to have important effects on health-seeking behavior on the continent. That is, irrespective of the low level of education in much of the general public in Africa, health-related education campaigns can have substantial impacts. Some successful examples are in HIV disease (26), polio (27, 28), and guinea worm (29).

One fact that our participants did not learn from our media was the name of the causative agent of KS — HHV-8/KSHV. This is unsurprising given the technical nature of this term and the low level of education we found in our participants. We do not, however, feel that this is a shortcoming of our media in that knowledge of the causative organism for KS is unimportant for community residents. Instead, what is essential is, and what we were seeking to accomplish, is for the public to recognize KS, seek immediate biomedical attention, and request a biopsy for diagnosis. We also paradoxically observed some participants changing their answer from a correct to incorrect one. We attribute this to uncertainty and guessing on the pre-test followed by incomplete learning and continued guessing on the post-test.

KS in Africa is similar to melanoma in other parts of the world (e.g., the U.S., Europe, and Australia). Like KS, melanoma clinically features hyperpigmented skin lesions (30) and has more favorable survival if diagnosed early (31). In the U.S., for example, the most current 5-year melanoma survival is >98% for those presenting with lesions of limited depth of invasion (32). Following documentation of increasing incidence and associated high mortality, public awareness campaigns for the early detection of melanoma, utilizing radio, television, and print media, begun in the 1970s in Australia (33), the 1980s in the United States (34, 35), and Europe (36, 37). The campaigns are ongoing and focus on the importance of routine skin self-examination for the appearance of new or changing hyperpigmented skin lesions and immediate presentation of skin changes to healthcare providers. Although it is controversial as to whether any of the intervention packages aimed at promoting earlier melanoma diagnosis actually reduce the incidence of melanoma-related death (38-44), that they increase early detection (as evidenced by increased percentage of early-stage disease amongst incident cases) (33, 45-49) provides proof-of-concept for considering this for KS in Africa. Furthermore, the many lessons learned in evaluating the effectiveness of these campaigns for melanoma (43, 50-52) may inform how effectiveness should be assessed for any similar campaign regarding early detection for KS in Africa. We note that exactly how an intervention campaign for KS in Africa should be constituted requires considerably more research, but we hypothesize that it would not be feasible to have routine comprehensive skin examination by healthcare providers. Instead, we speculate that the only feasible and most cost-efficient plan is a “bottom-up” approach focusing mainly on educating the public to be aware of KS, examine themselves and their loved ones, present immediately to healthcare providers if abnormalities are seen, and request biopsies for definitive diagnosis.

The magnitude of learning that our participants exhibited was admittedly modest. We attribute this to their receiving only a single exposure of one media type. In addition, even in those participants who did exhibit correct changes in their test performance after media exposure, it is unknown how long knowledge will be retained from this single exposure and, as noted above, whether it will translate into the desired behavior change. It is not our intention, however, to suggest that a single exposure to one media form will be impactful. Instead, we strongly believe that to cause a meaningful change in behavior, people will need to experience multiple exposures to multiple media types over an extended period of time. Directed discussions of the messages, for example, led by Village Health Team members (community health workers mandated with providing health education in the communities as well as linking to health services (53)), might add value to a public education campaign.

Our work does have limitations. While commercial markets are known to attract all types of individuals, we cannot be certain how our participants represent the overall general target population in Uganda. To mitigate this, we sampled markets in both urban and rural locations, and, furthermore, our participants’ literacy and HIV infection status mirrored national averages. Because we sought to evaluate learning from exposure to the media irrespective of reading literacy, our study staff both read the comics to those who were illiterate and administered the tests to all participants. It is therefore conceivable that this interaction with staff may have spuriously influenced test performance. To prevent this, staff were instructed to only read the text of the comics, point to the accompanying pictures, and not provide additional explanations. Pre- and post-tests followed a standardized script with no prompting. Finally, because the study was only conducted in Uganda, we are uncertain how findings will translate to other parts of Africa.

In conclusion, the high incidence and poor survival of KS in Africa — mainly due to advanced stage of disease at diagnosis — are a strong motivation to educate the public about KS and early detection. Although some will argue that a decline in KS incidence with growing ART availability (54, 55) lessens urgency, the incidence of KS in Africa shows no evidence of ever becoming lower than that of melanoma in resource-replete settings, and yet these regions continue to expend substantial resources in secondary prevention of melanoma. In Uganda, we have developed an educational message and several modes of delivery using common media that can be used as a foundation for educating the public about early KS detection.

### Data Availability

The data used to support the findings of this study may be released upon application to the Uganda National Council of Science and Technology, who can be contacted at.

## Acknowledgements

We are thankful to Straight Talk Foundation, Uganda and Henry Craig for their role in creating the media; our study team: Jane Frances Nalubega (Field Team Leader), Samuel Obol (Study Administrator), and the research assistants; the market management committees and community mobilisers of Hoima, Kampala, and Wakiso districts who permitted us to work in their premises. We especially appreciate all the study participants.

